# Next-Generation Skin Cancer Detection Using Efficient Fuzzy Fusion of Genomic and Imaging Data

**DOI:** 10.64898/2026.06.05.26355024

**Authors:** Afiur Rahaman Molla, Arnab Maity, Suvajit Saha, Rupsha Bhattacharya, Aritra Chakraborty, Surama Biswas, Subhrapratim Nath

**Affiliations:** Meghnad Saha Institute of Technology, Behind Urbana Complex Near,Ruby General Hospital, Anandapur Rd, Uchhepota, Kolkata, West Bengal 700150; alumni of Meghnad Saha Institute of Technology

**Keywords:** Fuzzy Fusion, Gene Expression Analysis, Image Analysis, Multimodal Learning, Skin Cancer Classification

## Abstract

Skin cancer requires early detection for improved survival rates. Most existing methods rely on deep learning–based image classification, which is affected by visual similarity among lesions. Fewer studies use Gene Expression (GE) analysis, which captures molecular characteristics but lacks structural and visual details. To overcome limitations of individual modalities, this paper proposes a multimodal framework integrating dermoscopic images and GE profiles for skin cancer classification. EfficientNet and logistic regression are used for image-based analysis and genomic skin lesion profiling, respectively, followed by fuzzy rule-based decision systems to reduce uncertainty within individual modalities. Finally, fuzzy fusion combines predictions from both modalities using uncertainty-based weighting of classifier outputs. The experimental findings show that both the image-based and GE–based classification models individually achieved accuracies of nearly 92%. However, the integration of prediction results through the proposed fuzzy fusion strategy further enhanced the classification performance, achieving an overall accuracy of 94.25%. The results obtained outperform contemporary methods, highlighting the effectiveness of combining complementary multimodal information compared with single-modality approaches.

## I. INTRODUCTION

SKIN cancer is one of the most prevalent and rapidly expanding types of cancer in the world today. Although Melanoma (MEL), Basal Cell Carcinoma (BCC), and Squamous Cell Carcinoma (SCC) are common types of skin cancer, MEL is responsible for the majority of skin cancer deaths despite representing a relatively small proportion of cases. Therefore, early and accurate identification of skin cancer types is crucial for reducing mortality rates. However, the traditional method for diagnosing skin cancer relies on dermoscopic image analysis by expert dermatologists and clinicians, which can be time-consuming and may vary depending on clinical interpretation. In recent years, automated skin cancer detection using deep learning has shown significant potential in supporting diagnosis and decision-making in dermatology clinics, diagnostic centers, and hospitals [1].

Convolutional Neural Networks (CNNs), a deep learning architecture, have shown excellent diagnosis accuracy on common dermoscopic image datasets, highlighting their usefulness as an assistive tool for healthcare professionals. An automated skin cancer detection system using CNN is proposed by Hasan et al. (2019) to distinguish between benign and malignant MEL dermoscopic images. The authors created a deep learning pipeline in which input images are preprocessed, followed by convolution operations for feature extraction, pooling operations for dimensionality reduction, fully connected layers for classification learning, and finally binary classification for distinguishing between two classes [2]. Nahata and Singh (2020) created a multi-class classification and skin cancer detection method based on CNN. The study evaluated five pre-trained CNN architectures, namely, ResNet50, MobileNet, VGG16, Inception V3, and InceptionResNet V2 [3]. A custom deep CNN model for detecting and classifying four skin cancers was introduced by Junayed et al., (2021), where the authors employed convolutional layers with increased filter depths, batch normalization, dropout, and a softmax layer as an output mechanism [4]. Ain et al. analyzed four multi-tree Genetic Programming (GP) methods, in which multiple decision trees evolve collaboratively for automatic skin cancer image classification, with emphasis on their behavior and effectiveness in real-world diagnostic settings [5]. A multimodal neural network system developed by Lyakhov et al., (2022) for classifying pigmented skin lesions into ten clinically important categories. The authors used dermoscopic images and patient statistical metadata for classification, achieving an accuracy of 83.6% [6]. A multimodal deep learning framework named MDFNet (Multimode Data Fusion Diagnosis Network) is proposed in [7] by Chen et al., (2023) for skin cancer classification using dermoscopic skin images and structured patient clinical data. A two-stage ensemble framework for binary MEL skin cancer detection was developed by Ghosh et al. (2024) using deep feature embeddings and achieved 91.4% accuracy [8]. Ozdemir and Pacal (2025) propose a hybrid deep learning model based on ConvNeXtV2 and self-attention layers to enhance feature extraction and improve classification performance for various skin cancer classifications, achieving an accuracy of 93.4% [9]. Mohamed et al. (2025) and Karnwal et al. (2026) both advocate a multi-modal deep learning framework designed to enhance the accuracy of skin cancer detection by combining heterogeneous data sources [10-11]. Gálvez et al. (2018), in their work introduced an innovative multiclass classification pipeline to characterize skin cancer using gene expression profiling across heterogeneous Affymetrix and Illumina platforms [13]. As shown by Gálvez et al (2019), gene expression studies using the combination of microarray and RNA-seq data provide a robust molecular framework that can be used to characterize several skin pathologies that cannot easily be differentiated by clinical or visual criteria alone. The authors employed machine learning algorithms like SVM, KNN, and Naïve Bayes to classify discriminative genes [14]. Bhalla et al. (2019) proposed a comprehensive machine learning-based study for predicting skin cutaneous MEL progression by analyzing multi-omics profiles from 466 patients obtained from The Cancer Genome Atlas project [15]. A CNN-based diagnostic model to classify oral SCC and differentiate it from other SCC (located in the head/neck) was developed by Pratama et al. (2021) using high-dimensional gene expressions from RNA sequence data [16]. A multimodal fusion framework is developed by Ou et al. (2022) for skin lesion diagnosis using smartphone-acquired clinical images and patient metadata, and got 76.8% accuracy [17]. Liu et al. (2022) proposed a multimodal framework for detecting metastatic MEL by integrating long non-coding RNAs (lncRNAs), protein-coding gene expression data, and pathology images [18]. The application of deep learning techniques for cancer classification using gene expression data was explored by Gupta et al. (2022) [19].

The literature review shows that both image-based and GE-based approaches for detecting skin cancer have advanced considerably in recent years. Image-based deep learning models are good at extracting spatial and visual patterns from dermoscopic images, while GE-based models provide useful molecular and biological insights into the lesion characteristics. GE models, however, are limited by the lack of structural and visual information, and are faced with the additional problems posed by the high dimensionality of genomic data with limited sample availability. Alternatively, image-based models are good at recognizing patterns but might not be able to distinguish between similar-looking lesion types. The majority of current studies focus on analyzing one modality and therefore do not effectively combine these complementary sources of information. Consequently, the fusion of imaging and genomic modalities may offer deeper diagnostic insights and improve the general precision and resilience of skin cancer classification algorithms.

Here in this paper, a multimodal framework is proposed to integrate dermoscopic image analysis and lesion GE profiling through fuzzy fusion for improved skin cancer classification. A total of 2,239 dermoscopic images from the International Skin Imaging Collaboration (ISIC) 2019 dataset [20] and 87 gene expression profiles from the National Center for Biotechnology Information, Gene Expression Omnibus [21] are preprocessed and analyzed using EfficientNet and Logistic Regression–based image and GE pipelines, respectively. The top three predictions from each pipeline are refined through fuzzy decision systems, and the final outputs are integrated using a fuzzy fusion unit for enhanced skin cancer classification. As an outcome, both the image and GE pipeline achieved an accuracy of 92%. Compared with these individual modalities, the proposed integrated fuzzy fusion model demonstrated significant performance improvement by achieving values of 94.25% for accuracy. The fused multimodal framework not only surpassed the performance of the individual pipelines but also outperformed previously reported methods.

## II. Proposed Methodology

In this paper, we propose an innovative framework for skin lesion classification by incorporating both of the following data modalities: dermoscopic images and GE profiles, to enhance diagnostic accuracy. The overall dual-pipeline architecture of the proposed framework, comprising the image and GE pipelines (see Panel A), along with the operation of the fuzzy decision maker within each pipeline (see Panel B) and the integration of both pipeline outputs using the fuzzy fusion unit (see Panel C), are depicted in Fig. 1.

**Fig 1.**
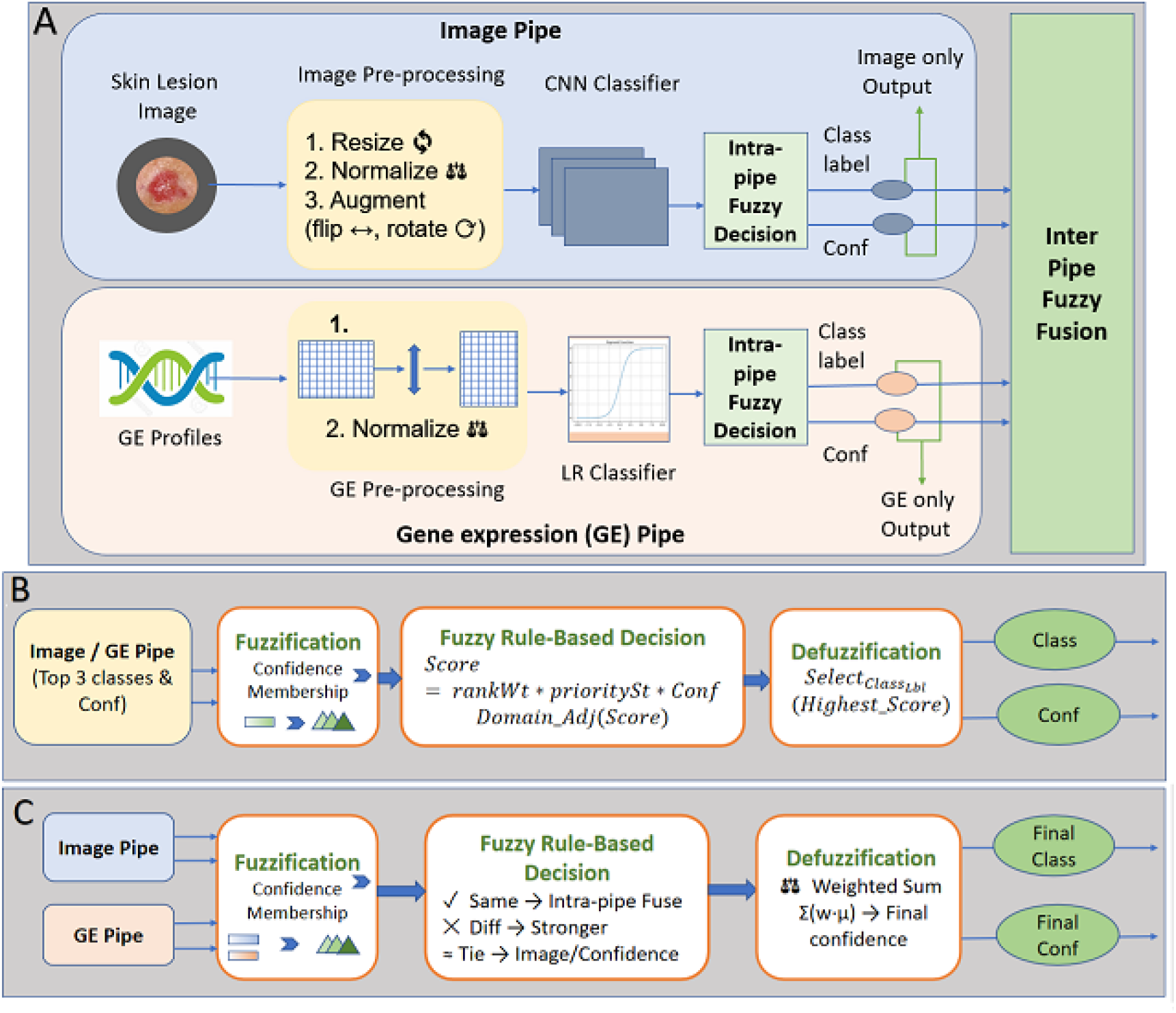
Overview of the Proposed Multimodal Framework: (A) Dual-Pipeline Architecture, (B) Intra-Pipe Fuzzy Decision, and (C) Inter-Pipe Fuzzy Fusion.

### A. Image Pipeline

For the image-based pipeline, skin lesion images are collected from a public dataset and subjected to preprocessing operations such as resizing, normalization, and data augmentation. Subsequent to the preprocessing phase, the discriminatory features of the dermoscopic images are acquired using the EfficientNet-B0 model for deep learning, whose backbone network is selected on account of its high efficiency.

The model is initialized with the pre-trained weights of EfficientNet, which allows it to benefit from transfer learning and achieve better generalization on unseen data. The input images first undergo preprocessing and data augmentation techniques. Augmentation techniques include resizing, normalization, and random transformations such as flipping and rotation to improve robustness and prevent overfitting. The preprocessed and augmented images are passed through the EfficientNet-B0 feature extractor. After extracting the features from input images, an average pooling layer is used. This pooling layer presents the summary of the feature maps by taking the average of each one and creating a dense vector representation of the whole image. A batch normalization has been applied to the outcome of the pooling layer. After the pooling layer, a dropout layer with a 50% dropout rate is applied to restrict overfitting. During training, dropout randomly deactivates half of the neurons, preventing the model from depending too much on specific features. Rather than memorizing the training data, this allows the network to acquire more robust and generalized patterns. An output layer is used to compute the probabilities (confidence score) of the skin lesion class, and the top three class-score pairs are fed to the Intra-Pipe Fuzzy Decision System (see subsection C of Section II) for the outcome of the image-pipe.

### B. Gene Expression (GE) Pipeline

The GE modality pipeline has been developed to identify genomic features that can be used for skin lesion classification using a statistical learning approach. At first, a dataset containing gene expression profiles of multiclass skin cancer patients vs. normal (NORM) is collected from a public database like NCBI, GEO [21]. To ensure data consistency and quality, the raw gene dataset is preprocessed.

Samples with missing gene symbols are removed. All probes representing the same gene are aggregated by calculating the mean expression values. After the aggregation, the matrix is transposed so that the rows represent samples and columns represent the genes. Class labels are assigned to each sample based on its corresponding categories. Afterward, the label encoding technique is used, where the categorical labels are converted into numerical form. This process allows the model to work efficiently with the output variable. To ensure uniform feature scaling, the GE values are then scaled using Min-Max normalization, which maps GE values into the range [0,1]. In this work, to avoid overfitting and reduce computational complexity, a new feature selection strategy is employed. Here, the SelectKbest with the ANOVA F-statistic is used to rank genes based on the separability among classes. Based on these scores, the 50 most relevant and informative genes are selected for model training. A logistic regression model is trained on the selected feature as the classifier for the GE pipeline. To address class imbalance, a balanced class weighting strategy is incorporated. The training and evaluation process for the model incorporates the 5-fold cross-validation technique to ensure robustness and prevent overfitting. The LR classifier outputs the probability scores for the classes of the samples, and among them, the top K scores (K=3) are chosen and then post-processed by using an intra-pipe fuzzy decision system to get the genomics prediction.

### C. Intra-Pipe Fuzzy Decision System

To lower classification uncertainty, an intra-pipe fuzzy decision system processes the top three projected class labels and their confidence scores within each pipeline. Triangular membership functions are used to fuzzify each confidence score into low, medium, and high membership categories. The fuzzy strength, prediction rank weight, and initial classifier confidence score are combined with domain-specific modifications that, where necessary, marginally favor alternative classes and punish uncertain dominant-class predictions to determine the final decision score. Here, the confidence score reflects the classifier’s initial prediction probability, the rank weight gives higher-ranked predictions more weight, and fuzzy strength indicates a prediction’s degree of certainty. The class that receives the highest fuzzy decision score is chosen to be the pipeline’s final output (see panel B of Fig. 1).

### D. Inter-Pipe Fuzzy Fusion

When the image and gene expression (GE) pipelines produce refined outputs, an inter-pipe fuzzy fusion mechanism is used to produce the final multimodal prediction. First, the confidence scores of both pipelines are fuzzified into low, medium, and high confidence levels by means of triangular membership functions. When the predictions of the two pipelines are identical, their confidence memberships are merged through a weighted fuzzy defuzzification process to produce a single fused confidence score, which eventually improves the reliability of the prediction using consensus-based integration. If the pipelines disagree on the class, the final decision is based on a comparison of the fuzzy confidence strengths of the pipelines, and the prediction with the stronger fuzzy significance is chosen. If both pipelines exhibit equal fuzzy strength and identical confidence values, preference is given to the image pipeline because of its stronger spatial representation capability. By integrating complementary visual and molecular information while systematically handling uncertainty, the proposed inter-pipe fuzzy fusion strategy improves the robustness and accuracy of the final skin cancer classification output.

### E. Metrics of Performance

The Performance of the proposed imaged, GE and fused modality pipeline has been evaluated using five quantitative measures, including precision ( *PR*), recall (*RE*), F1-score (*F*1), Accuracy, and an important quantity called False Positive Rate (FPR) defined as follows:

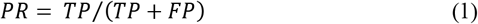

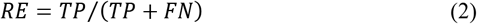

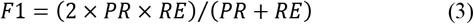

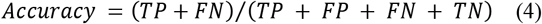

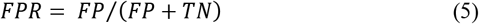

Here, *TP, TN, FP*, and *FN* stand for true positives, true negatives, false positives, and false negatives, respectively.

## IV. Result and discussion

This paper investigates the performance of fuzzy fusion techniques using image, GE, and multimodal approaches for skin cancer classification. To evaluate the performance of the individual pipelines and the suggested integrated framework in terms of calculating *PR*(Eq. 1), *RE* (Eq. 2), *F*1 (Eq. 3), and classification accuracy (Eq. 4), we have performed several experiments. The tests are conducted using a Tesla T4 GPU runtime, 12.7 GB of RAM, and 78 GB of free disc space on the cloud-based platform Google Colab. The primary deep learning and machine learning frameworks used in the Python 3.10 programming environment are TensorFlow 2.x and Scikit Learn, respectively.

### A. Performance Evaluation of the Image Pipeline

The image dataset employed here is collected from ISIC 2019 [20]. It contains nine skin lesion classes with an almost balanced sample distribution. The number of samples ranges from 454 in Actinic keratosis (AK) to 561 in BCC, and the remaining classes are distributed between 480 and 560 samples (see Supplement 1). An unbiased nature (not biased to any particular class) of the model is ensured by this balanced distribution, which improves evaluation reliability. The dataset is divided into 80% and 20% for training and testing, respectively, where the training set optimizes model parameters and the test set evaluates generalization performance. After training, the test image data is evaluated using the Image Pipeline, explained in Subsection A of Section II.

Table 1 shows the per-class accuracy report, containing columns like #Image (number of images per class), and performance measures like *PR, RE*, and *F*1, providing an in-depth analysis of the classification model’s performance across all nine classes. Dermatofibroma (DF) and vascular lesion (VL) achieved perfect accuracy of 1.00, while SCC, pigmented benign keratosis (PBK), and BCC achieved accuracies of 0.98, 0.97, and 0.96, respectively, indicating near-perfect classification. Seborrheic keratosis (SK) and AK exhibited moderate accuracies of approximately 0.88 and 0.86, respectively. However, MEL and nevus (NV) show lower accuracies of 0.79 and 0.85, respectively, due to their strong visual similarities; as a result, the final overall accuracy (average of individual class accuracies, calculated by Eq. 4) recorded as 91.99% ≈ 92%.

**Table 1.** Per-class Classification Report on Image Data.

| Class | #Image | $PR$ | $RE$ | $F1$ |
| --- | --- | --- | --- | --- |
| AK | 454 | 0.85 | 0.86 | 0.86 |
| BCC | 561 | 0.99 | 0.96 | 0.98 |
| DF | 480 | 0.99 | 1.00 | 0.99 |
| MEL | 553 | 0.78 | 0.79 | 0.79 |
| NV | 517 | 0.89 | 0.85 | 0.87 |
| PBK | 560 | 0.96 | 0.97 | 0.96 |
| SK | 539 | 0.99 | 0.98 | 0.98 |
| SCC | 517 | 0.83 | 0.87 | 0.85 |
| VL | 514 | 1.00 | 1.00 | 1.00 |
| **Overall Accuracy** | **0.9199** |  |  |  |

The model’s performance across all nine classes is shown in the confusion matrix in Fig. 2, with the majority of images being assigned to the diagonal portion. DF, VL, and SCC recorded the fewest misclassifications with 478, 513, and 526 correct predictions, respectively. MEL is identified as the most challenging class to classify since there are 84 cases of MEL that are misclassified as SK and 30 cases as NV. Additionally, for NV and AK, a significant number of cases are misclassified. The distribution of *F*1 of different classes (see Supplement 2) is computed with Eq. 3.

**Fig 2.**
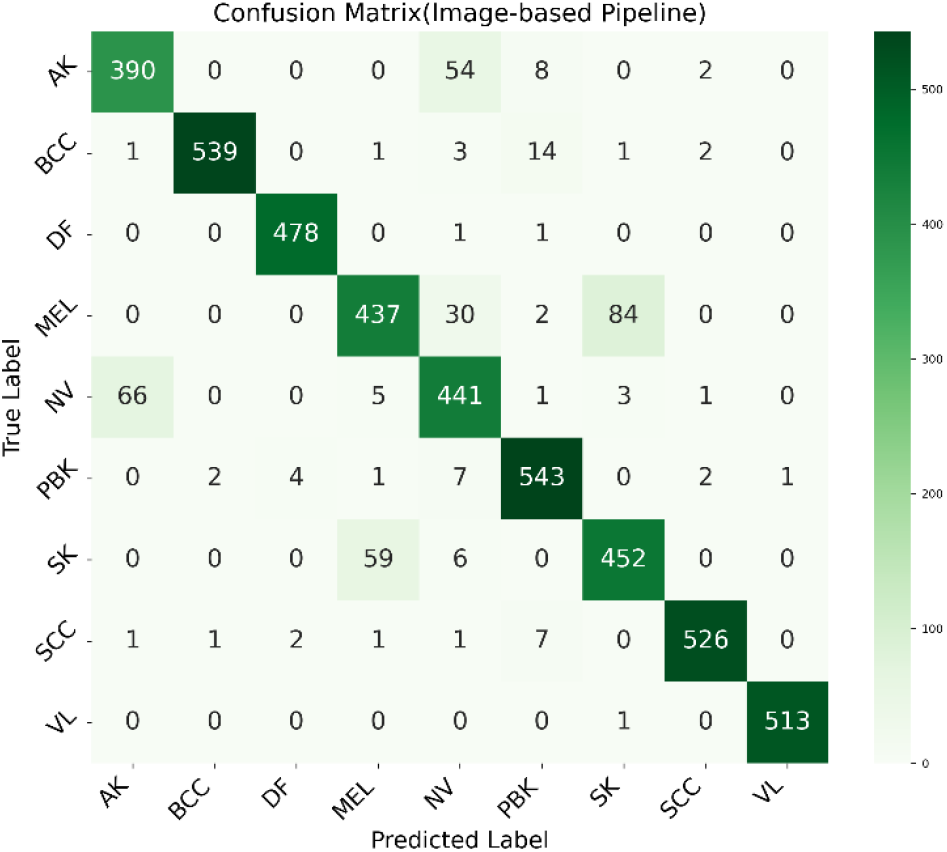
Confusion Matrix of the Image-based classification model

### B. Performance Evaluation of GE Pipeline

A multiclass skin cancer gene expression dataset (GSE7553) is obtained from the NCBI, GEO and subjected to the preprocessing procedures described in Subsection B of Section II. The performance of the GE pipeline is evaluated using a balanced logistic regression classifier. To verify model stability and consistency across folds during training/validation, a stratified 5-fold cross-validation technique is used. Fold-wise accuracy and aggregated accuracy scores are provided in Table 2. With a gradual increase across folds, folds 4 and 5 exhibit the highest accuracy (1.00), which indicates that the model can achieve complete generalization if the training folds represent all the classes equally. The comparatively lower accuracy in fold 1 can be explained by the small counts of NORM (n=5) and SCC (n=11) classes, which introduces stochastic variability in stratified splitting at this dataset scale.

**Table 2.** Fold-Wise Cross-Validation Accuracy of the LR Classifier.

| Fold 1 | Fold 2 | Fold 3 | Fold 4 | Fold 5 | Mean ± SD |
| --- | --- | --- | --- | --- | --- |
| 0.722 | 0.944 | 0.941 | 1.000 | 1.000 | 0.922 ± 0.110 |

From the evaluation of the GE pipeline using a Logistic Regression classifier, a detailed per-class classification report is presented in Table 3. It is observed that BCC and MEL exhibit superior performance metrics with *F*1 of 0.97 and 0.95, respectively. The model achieved an overall accuracy of 92.16% ≈ 92%, demonstrating a high level of discriminatory power among the selected GE patterns.

**Table 3.** Per-class classification report of GE Pipe.

| Class | #Sample | PR | RE | F1 |
| --- | --- | --- | --- | --- |
| BCC | 15 | 1.00 | 0.93 | 0.97 |
| MEL | 56 | 0.98 | 0.93 | 0.95 |
| NORM | 5 | 0.50 | 0.80 | 0.62 |
| SCC | 11 | 0.83 | 0.91 | 0.87 |
| **Overall Accuracy** | **0.9216** |  |  |  |

Fig. 3 represents the confusion matrix derived from 5-fold cross-validation. BCC is correctly identified in 14 of 15 cases, with 1 sample misclassified as SCC. MEL is correctly identified in 52 of 56 cases, with 3 samples misclassified as NORM and 1 as SCC. SCC is correctly identified in 10 of 11 cases, with one sample assigned to NORM. NORM is correctly identified in 4 of 5 cases, with one sample assigned to MEL (Supplement 3 contains the training validation curve).

**Fig 3.**
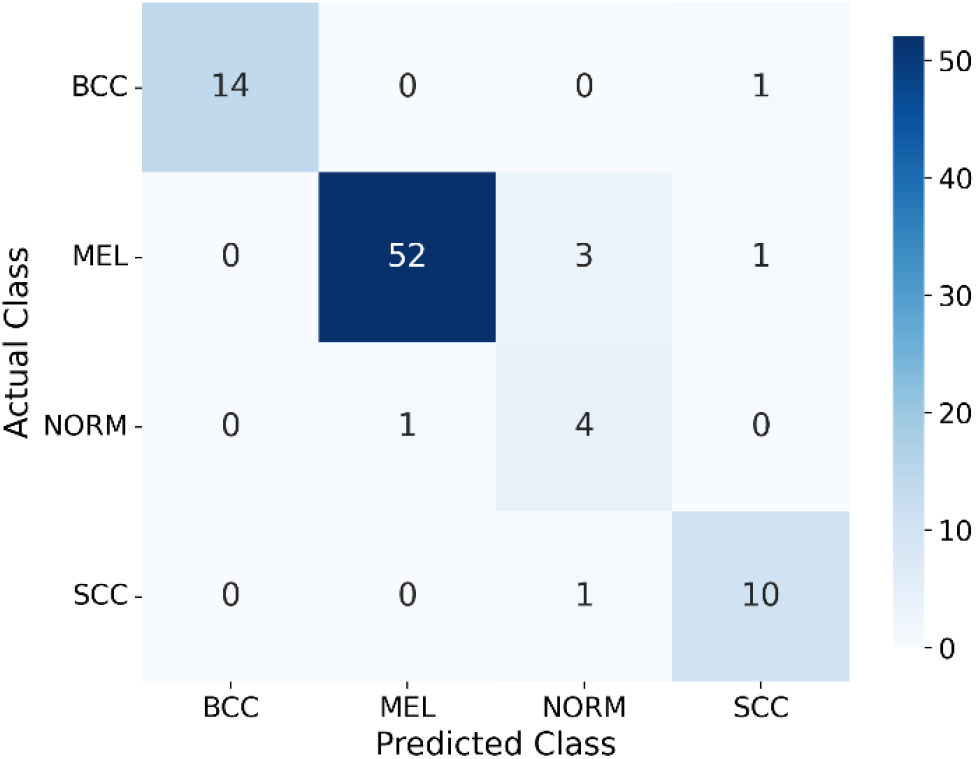
Confusion matrix of GE Pipe

Strong classification performance is shown by the Receiver Operating Characteristic (ROC) analysis, which computes the relation between the true positive rate or *RE* (Eq. 2) and *FPR*calculated by Eq. 5 across various classification thresholds. As shown in Table 4, the matching Area under the Curve (AUC) values, which show how well the model can differentiate between various classes, are found to be high for the majority of classes. In this case, SCC achieved 0.99 and MEL obtained 0.96 of AUC values, while BCC earned an AUC of 1.00. The ROC curves may be observed in Supplement 4.

**Table 4.** Per-class ROC-AUC Analysis of GE Pipe.

| BCC | MEL | NORM | SCC |
| --- | --- | --- | --- |
| 1.00 | 0.96 | 0.84 | 0.99 |

### C. Performance Evaluation of Inter-Pipe Fuzzy Fusion

To utilize the complementary information provided by both the image and GE modalities, an inter-pipe fuzzy fusion method (see Subsection E of Section II) is employed to integrate the predictions made by the individual models. The proposed fusion approach achieved an overall accuracy of 94.25%, outperforming both the image and GE pipeline.

A detailed per-class classification report for the common classes available in both the Image and GE pipelines of the proposed fusion model is presented in Table 5. A notable performance improvement has been observed in BCC and SCC with a *PR, RE*, and *F*1 of 1.00. MEL, on the other hand, maintains strong performance with a *PR* of 0.98 and an *F*1 of 0.95, highlighting the model’s ability to accurately identify this clinically critical class. NORM classification remains consistent, with *PR* 0.50, *RE* 0.80, and *F*1 0.62, likely due to overlapping feature characteristics and limited sample representation. The bar chart based on this matrix is available in Supplement 5. Fig. 4 shows the confusion matrix of the proposed fusion model for skin lesion classification, where all BCC and SCC cases are correctly identified.

**Table 5.** Per-Class Classification Report of the Fuzzy Fusion Model.

| Class | PR | RE | F1 |
| --- | --- | --- | --- |
| BCC | 1.00 | 1.00 | 1.00 |
| MEL | 0.98 | 0.93 | 0.95 |
| Normal Skin | 0.50 | 0.80 | 0.62 |
| SCC | 1.00 | 1.00 | 1.00 |
| **Overall Accuracy** | **0.9425** |  |  |

**Fig 4.**
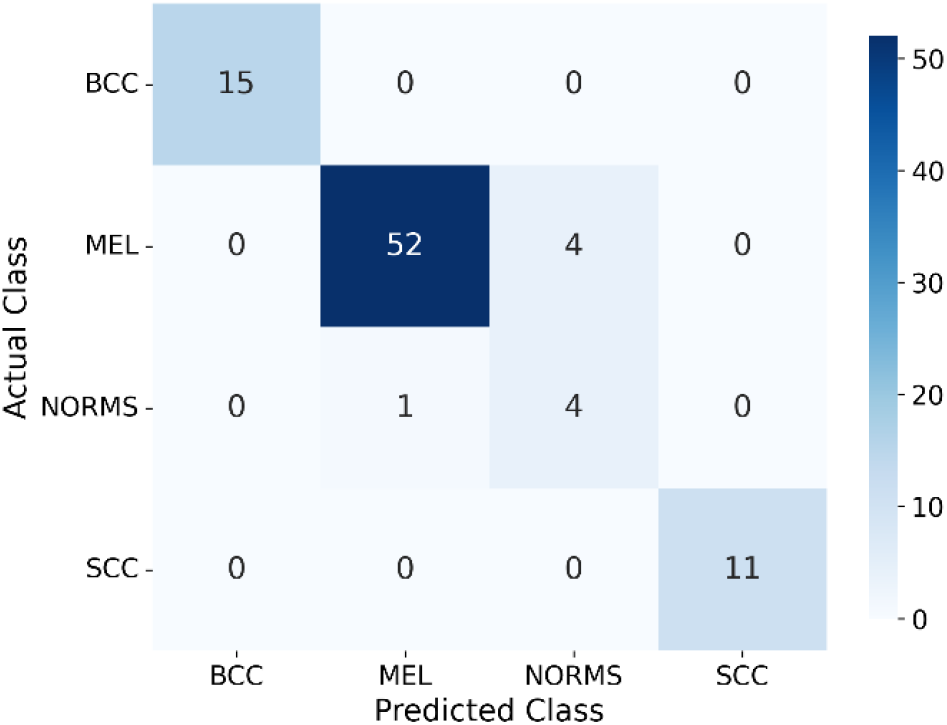
Performance Evaluation of the Multimodal Fusion Model

Fig. 5 represents a comparative analysis of the proposed model against several existing approaches. The proposed method achieves an accuracy of 94.25%, significantly outperforming previously reported methods, including Ozdemir and Pacal (93.4%), Ghosh et al. (91.6%), Chen et al. (83.5%), and Lyakhov et al. (83.6%). In conclusion, the experimental findings show that, despite certain restrictions, both the image-based and GE-based models exhibit good individual performance. However, through the introduction of the multimodal fusion strategy, the limitations have been addressed, resulting in better accuracy levels.

**Fig 5.**
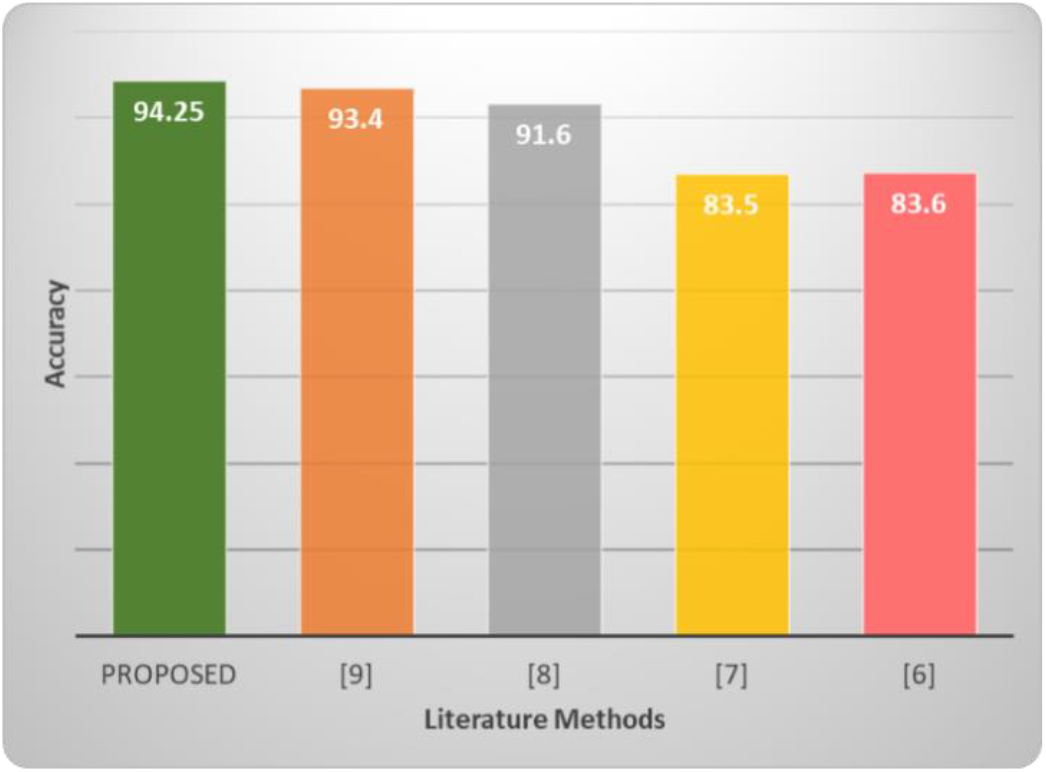
Accuracy Comparison with Existing Methods.

## V. Conclusion

This paper presented a multimodal skin cancer classification framework that integrates dermoscopic image analysis and lesion and gene expression profiling through a fuzzy fusion mechanism. By combining the complementary strengths of EfficientNet-based image analysis and Logistic Regression– based gene expression classification, the proposed framework effectively addressed the limitations associated with single-modality approaches. The incorporation of intra-pipe fuzzy decision and inter-pipe fuzzy fusion reduced uncertainty and improved the reliability of classification outcomes. Experimental evaluation demonstrated that the integrated multimodal framework significantly outperformed the individual image and gene expression pipelines in terms of accuracy, *PR, RE*, and *F*1. The obtained results also surpassed several existing literature methods, highlighting the effectiveness of combining visual and molecular lesion information for accurate skin cancer detection.

## Data Availability

All data produced in the present study are available upon reasonable request to the corresponding author.

## References

[1] M. L. Wei, M. Tada, A. So, and R. Torres, “Artificial intelligence and skin cancer,” Front. Med., vol. 11, pp. 1331895, 2024.

[2] M. Hasan, S. D. Barman, S. Islam, and A. W. Reza, “Skin cancer detection using a convolutional neural network,” in Proc. 5th Int. Conf. Computing and Artificial Intelligence (ICCAI), Apr. 2019, pp. 254–258.

[3] H. Nahata and S. P. Singh, “Deep learning solutions for skin cancer detection and diagnosis,” in Machine Learning with Health Care Perspective: Machine Learning and Healthcare, Cham, Switzerland: Springer International Publishing, 2020, pp. 159–182.

[4] M. S. Junayed, N. Anjum, A. Noman, and B. Islam, “A deep CNN model for skin cancer detection and classification,” Comput. Sci. Res. Notes, vol. 3101, pp. 71–80, 2021.

[5] Q. U. Ain, H. Al-Sahaf, B. Xue, and M. Zhang, “Automatically diagnosing skin cancers from multimodality images using two-stage genetic programming,” IEEE Trans. Cybern., vol. 53, no. 5, pp. 2727–2740, 2022.

[6] P. A. Lyakhov, U. A. Lyakhova, and N. N. Nagornov, “System for the recognizing of pigmented skin lesions with fusion and analysis of heterogeneous data based on a multimodal neural network,” Cancers, vol. 14, no. 7, pp. 1819, 2022.

[7] Q. Chen, M. Li, C. Chen, P. Zhou, X. Lv, and C. Chen, “MDFNet: Application of multimodal fusion method based on skin image and clinical data to skin cancer classification,” J. Cancer Res. Clin. Oncol., vol. 149, no. 7, pp. 3287–3299, 2023.

[8] S. Ghosh, S. Dhar, R. Yoddha, S. Kumar, A. K. Thakur, and N. D. Jana, “MEL skin cancer detection using ensemble of machine learning models considering deep feature embeddings,” Procedia Comput. Sci., vol. 235, pp. 3007–3015, 2024.

[9] B. Ozdemir and I. Pacal, “A robust deep learning framework for multiclass skin cancer classification,” Scientific Reports, vol. 15, no. 1, pp. 4938, 2025.

[10] E. H. Mohamed, N. Abdu, M. Khalil, H. Kamal, and E. A. Rashed, “MiSC: A hybrid multi-modal deep learning approach for accurate skin cancer detection,” Multimedia Tools Appl., vol. 84, no. 36, pp. 45321–45345, 2025.

[11] A. Karnwal, M. Selvaraj, G. Kumar, A. Kumar, A. R. M. S. Al-Tawaha, and N. Nesterova, “Multimodal artificial intelligence for enhanced skin cancer diagnosis and prognosis,” Discover Oncology, 2026.

[12] H. Mitsui et al., “Gene Expression profiling of the leading edge of cutaneous squamous cell carcinoma: IL-24-driven MMP-7,” J. Investigative Dermatology, vol. 134, no. 5, pp. 1418–1427, 2014.

[13] J. M. Gálvez et al., “Multiclass classification for skin cancer profiling based on the integration of heterogeneous GE series,” PLoS One, vol. 13, no. 5, p. e0196836, 2018.

[14] J. M. Galvez et al., “Towards improving skin cancer diagnosis by integrating microarray and RNA-seq datasets,” IEEE J. Biomed. Health Informatics, vol. 24, no. 7, pp. 2119–2130, 2019.

[15] S. Bhalla, H. Kaur, A. Dhall, and G. P. S. Raghava, “Prediction and analysis of skin cancer progression using genomics profiles of patients,” Sci. Rep., vol. 9, no. 1, p. 15790, 2019.

[16] R. Pratama, J. J. Hwang, J. H. Lee, G. Song, and H. R. Park, “Authentication of differential GE in oral squamous cell carcinoma using machine learning applications,” BMC Oral Health, vol. 21, no. 1, p. 281, 2021.

[17] C. Ou et al., “A deep learning based multimodal fusion model for skin lesion diagnosis using smartphone collected clinical images and metadata,” Front. Surg., vol. 9, p. 1029991, 2022.

[18] S. Liu, Y. Fan, K. Li, H. Zhang, X. Wang, R. Ju, Y. Zhang, and F. Zhou, “Integration of lncRNAs, protein-coding genes and pathology images for detecting metastatic MEL,” Genes, vol. 13, no. 10, p. 1916, 2022.

[19] S. Gupta, M. K. Gupta, M. Shabaz, and A. Sharma, “Deep learning techniques for cancer classification using microarray GE data,” Front. Physiol., vol. 13, p. 952709, 2022.

[20] M. Combalia, N. Codella, V. Rotemberg, C. Carrera, S. Dusza, D. Gutman, et al., “Validation of artificial intelligence prediction models for skin cancer diagnosis using dermoscopy images: The 2019 International Skin Imaging Collaboration Grand Challenge,” The Lancet Digital Health, vol. 4, no. 5, pp. e330–e339, 2022.

[21] E. Clough, T. Barrett, S. E. Wilhite, P. Ledoux, C. Evangelista, I. F. Kim, et al., “NCBI GEO: archive for gene expression and epigenomics data sets: 23-year update,” Nucleic Acids Research, vol. 52, no. D1, pp. D138–D144, 2023.

